# COVID-19 in India: Predictions, Reproduction Number and Public Health Preparedness

**DOI:** 10.1101/2020.04.09.20059261

**Authors:** Balram Rai, Anandi Shukla, Laxmi Kant Dwivedi

## Abstract

**Introduction:** The COVID-19 has emerged as a global concern for public health due to large scale outbreak. The number of confirmed cases has also been increased in India in past few weeks. The predictions for the COVID-19 can provide insights into the epidemiology of the disease, which helps policymakers to check health system capacities.

**Methods:** We obtained data on daily confirmed, recovered and deaths cases for a period of 21 days and have implemented the exponential growth model to predict the future cases for all the three components. The mathematical model was used to calculate the average reproduction number and herd immunity. We estimated the number of active cases till 30^th^ of April. We have also tried to analyze the public health capacity to combat COVID-19 in India.

**Results:** If the exponential growth in number of cases continue then the total number of active cases will be 2,49,635 until the end of April. The reproduction number for COVID-19 in India was found to be 2.56 and herd immunity as 61%. The cumulative cases predicted by the mathematical model was 1,20,203.

**Discussion:** This prediction provides an alarming situation for India in terms of public health preparedness. The number of tests is needed to increase to detect all the cases of COVID-19 in India. Though some serious preventive measures have been implemented, but India should be ready to face any sudden community outbreak.

## Introduction

The cases of novel coronavirus (COVID-2019)-infected pneumonia started since the 19th of December, 2019, in Wuhan (Central China). A large scale outbreak of the disease resulted in a pandemic. It became a public health concern all over the world.^1^ On the 7th of January, 2020, the etiological agent of the outbreak was identified as a novel coronavirus^2^, and it was renamed as COVID-19 by WHO on the 12th of February, 2020. It is a zoonotic coronavirus which is similar to SARS and MERS coronavirus.^3^

The WHO announced it as a “Public health emergency of international concern” on the 30th of January,2020.^4^ As per the report published by the World Health Organization on the 4th of April, there are 1051635 confirmed cases, around 57 thousand deaths due to COVID-19. About 200 countries, areas, and territories are there with cases of COVID-19 in the world.^5^ In much of the world, people exhibiting mild or no symptoms are unable to get tested, meaning that the actual number of cases could be much higher.^6^ In the past few weeks, countries such as Italy, Spain, and the United States have shown a massive increase in the number of cases and deaths per day.^7^ Even after the gradual increase in facilities, the situations in these countries are still quite concerning.^8^

In India, 17% of the world population resides in 2.4% area of the world, resulting in a densely populated country, which makes it a high-risk country for any infectious disease. The study of various countries has shown that the explosion of the COVID-19 disease can be seen after a few days. In that case, India will not be an exception, and it will be wrong to state that the corona crisis will not impact India.

In India, the first positive case of the COVID-19 was detected on the 30th of January, 2020, in Kerala.^9^ The cases started to increase from the first week of March 2020. The majority of the patients initially identified had a traveling history. These cases worked as primary cases and infected others. Hence, it becomes a thing of utmost importance to estimate the transmission dynamics in the initial days of infectious disease outbreak and inform predictions about the potential growth of cases.^10^ This prediction can provide insights into the epidemiology of the disease, which helps policymakers to check health system capacities. It can identify whether the control measures are having a measurable effect or not^11-12^; also, a prediction model can be updated to estimate the risk for other countries.^13^ The transmissibility of COVID-19 from human to human is sufficient to support sustained transmission unless certain ontrol measures are implemented.^14^

### Data and Methodology

Data on COVID-19 was taken from the data-sharing portal covid19india.org, which was also verified by the Ministry of Health and Family Welfare of India and health ministries of different states. We have collected information on three main components of interest, i.e., daily confirmed cases, daily recovered cases, and daily deaths from 14^th^ of March to 3^rd^ of April 2020 for a period of total 21 days. We emphasized the active cases of COVID-19, which has been adjusted by recoveries and deaths. We also calculated the cumulative number of cases for all the three components presented in figure 1.

### Exponential Growth model

Since we have data on cumulative confirmed cases, recovered cases, and deaths for each day for this period of 21 days, it is possible to fit an exponential growth model for these three data patterns.

The basic exponential growth model is described as 

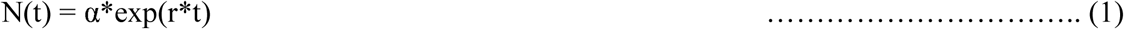

Where N(t) = number of cases at any time ‘t.’

α = constant

r = growth rate

t = time (day)

This growth model was adopted for all three components and predicted the future cases up to the end of the month, April, i.e., the 30th of April. While all three components closely followed exponential growth with a rise in the number of confirmed cases more steadily compared to recovered cases and deaths. The exponential growth rate model for confirmed cases, recovered cases, and deaths are based on the available data is described in equations 2, 3, and 4, respectively. The active cases were calculated by subtracting the total recoveries and deaths from total confirmed cases described in equation 5. The active cases refer to the cases that will be active at that time and can require hospitalization and ICU (Intensive care unit) in case of more severity. 

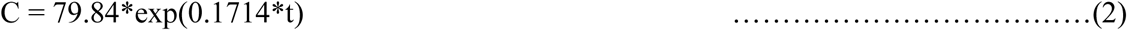

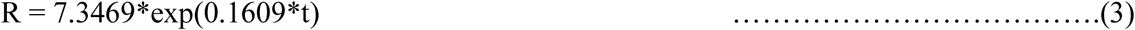

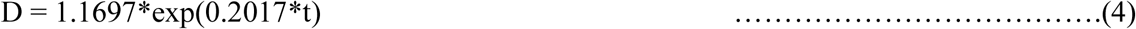

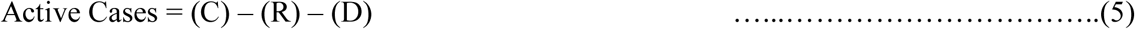

The number of cases obtained from the above growth models is consistent with the cases observed from the data used, which can be reflected in figure 1.A, 1.B, and 1.C. Hence these growth models can provide us a fairly enough estimates which can be useful for the policymakers to be prepared for any such public health requirements for hospitalizations and ICU.

**Figure 1.A.**
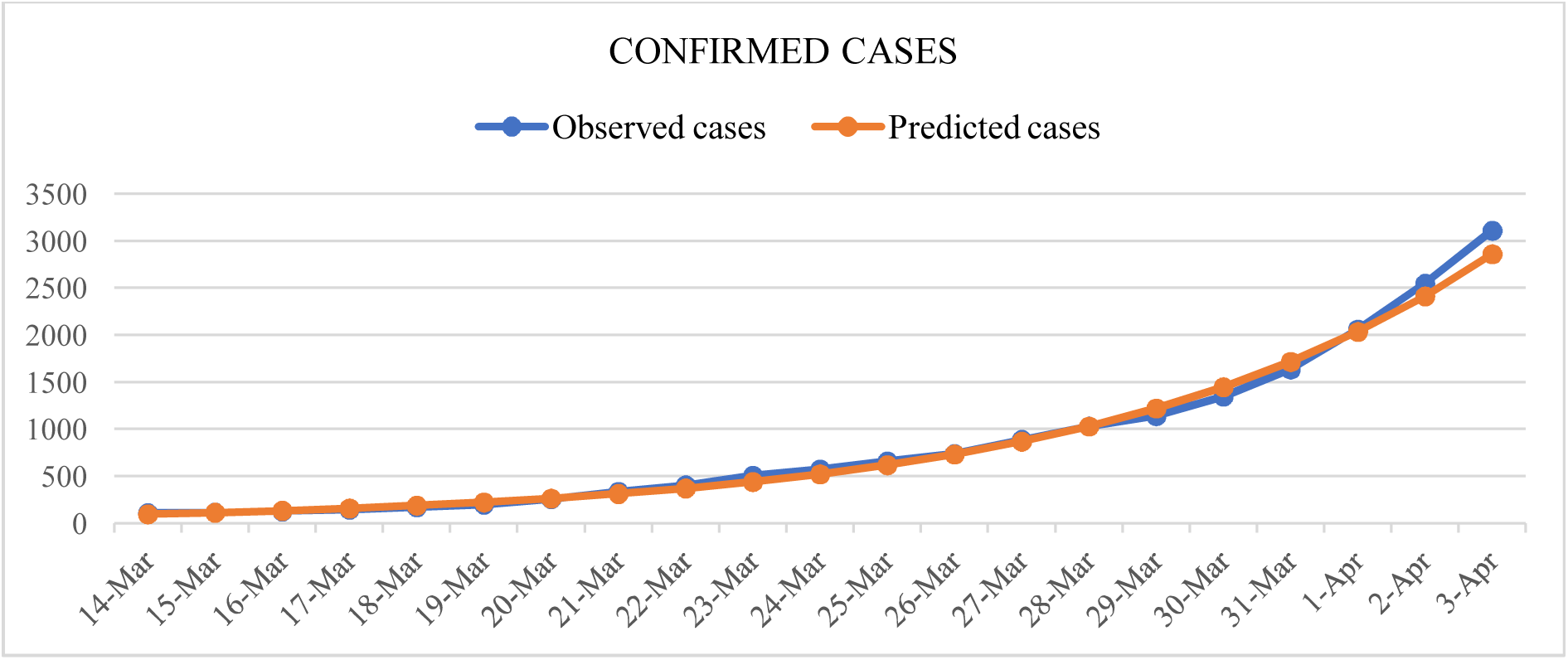
Daily cumulative confirmed cases based on exponential growth model from COVID-19 in India

**Figure 1.B.**
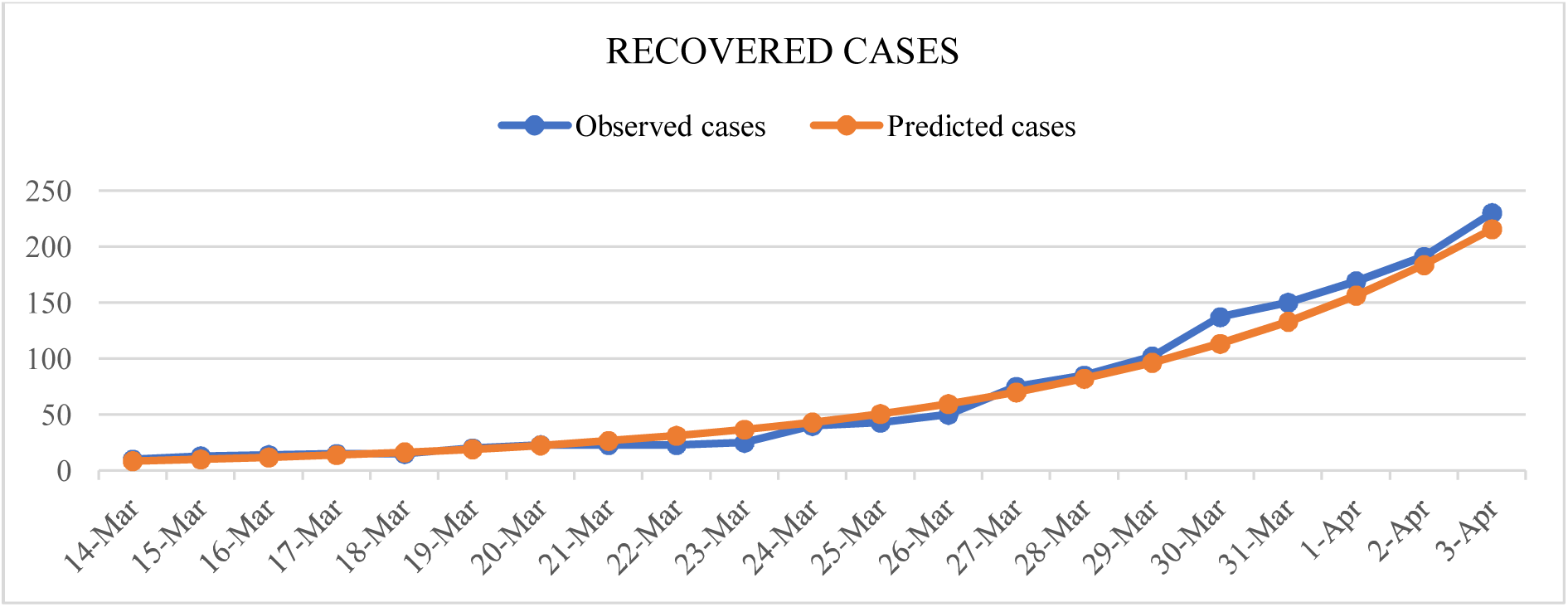
Daily cumulative recovered cases based on exponential growth model from COVID-19 in India

**Figure 1.C.**
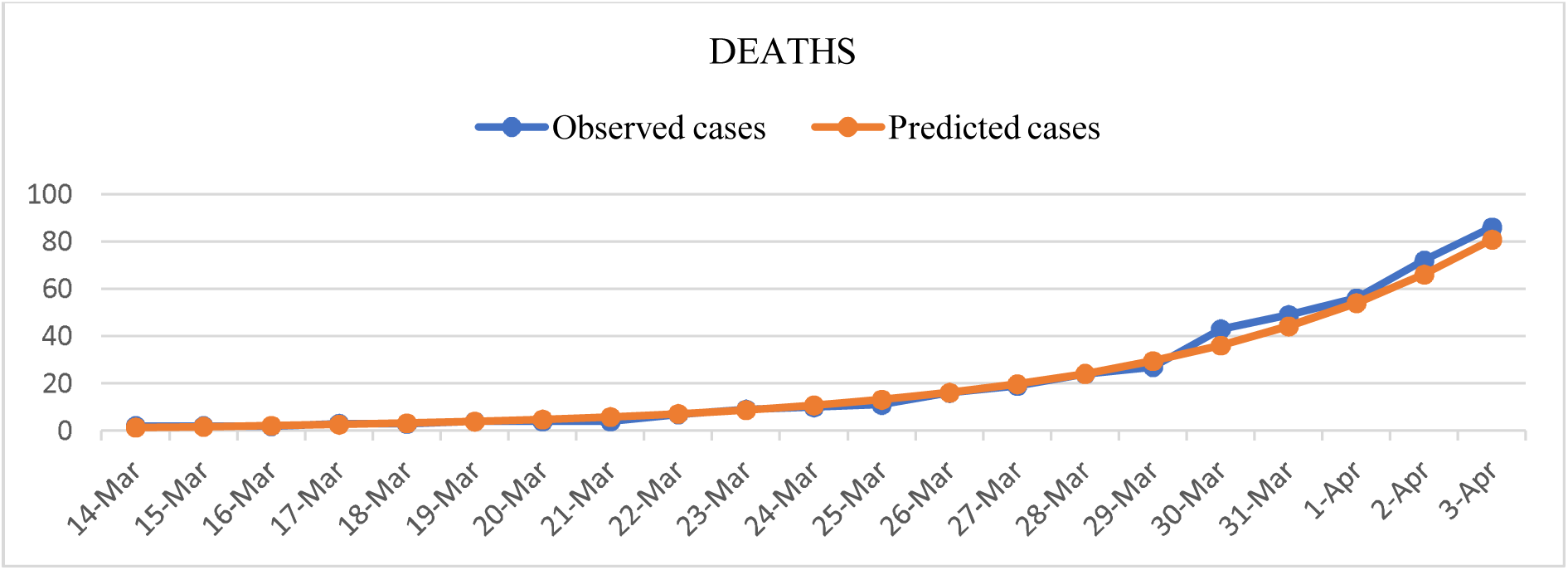
Daily cumulative deaths based on exponential growth model from COVID-19 in India

### Mathematical Model for reproduction number and Herd Immunity

The mathematical model for calculating the number of incidence cases based on reproduction number is defined as : 

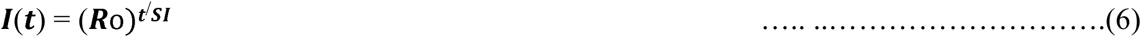

I(t) = Number of incidence cases at time t

Ro = Reproduction number

SI = Serial interval t = Prediction time

The number of cumulative cases have been calculated based on this mathematical model until the end of April.

### Estimation of Reproduction number and Herd Immunity

The reproduction number is defined as the average number of new infections generated by one infected individual during the entire infectious period in a fully suspectible population. The basic reproduction number reflects the ability of infection spreading the infectious period under no control.^15^ The approach used to estimate the basic reproduction number in this model is to calculate the average Ro based on the daily cumulative cases for 21 days described in equation 7. The herd immunity(HI) is estimated based on reproduction number by equation 8. The herd immunity basically indicates the resistance to the spread of a infectious disease within a population that results if a sufficiently high proportion of individuals are immune to the disease, especially through vaccination. 

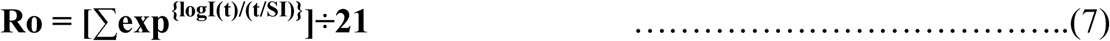

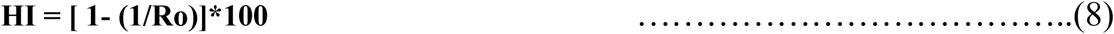

Serial Interal is the time between onset of a primary and secondary case. Due to unavailability of detailed data on this parameter in we have used SI as 4.4 days as reported in previous studies.^16^

### Public health capacity of India

To ensure the public health preparedness of India we have collected information on different public health facilities available in India including number of hospitals and beds available. Information on total government hospitals and beds as on July 2019 have been taken from the Open Govermnent data source provided by Ministry of Health and family Welfare.^17^ To estimate the distribution of public health facilities among the population we have used the projected population of India for the year 2019 (Report of the Technical Group on Population Projections).^18^ The total projected population of India for the year 2019 is 133,29,00,000. The burden on public health facilities due to COVID-19 is estimated by total number of cases of COVID-19 per hospital needs to accommodate if the number of cumulative cases is projected by exponential growth model and mathematical model on Ro.

## Results

Table 1 provides the prediction for confirmed cases, recovered cases, and deaths for COVID-19 in India till the 30th of April. If the increase in the number of cases continues to grow exponentially in upcoming days, then the number of active cases will rise to 2,49,365, and death toll will rise to 18,739. The number of recovered cases is expected to be 16,605 until the end of April. The number of confirmed cases will reach to 50,000 and then 1,00,000 on the 20th of April and the 24th of April, respectively. The exponential growth models fit very closely with the number of observed cases that indicates India has already reached the exponential phase of growth in the number of COVID-19 cases. Most countries have experienced exponential growth in COVID-19 cases, including China, USA, Italy, Spain, Germany, France. It took only 15 days in the USA to climb the number of confirmed cases from 1,000 to 1,00,00 as reported by center for disease control and prevention.^19^ The exponential phase may continue to increase until some serious preventive measures are taken (Figure 2).

**Table 1.** Predictions for COVID-19 by exponential growth model in upcoming days in India until 30^th^ April,2020

|  |  |  |  |  |
| --- | --- | --- | --- | --- |
| Date | Confirmed Cases | Recovered cases | Deaths | Active cases |
| 04-April | 3391 | 253 | 99 | 3039 |
| 05-April | 4021 | 297 | 121 | 3602 |
| 06-April | 4768 | 349 | 148 | 4270 |
| 07-April | 5653 | 410 | 181 | 5062 |
| 08-April | 6704 | 482 | 222 | 6000 |
| 09-April | 7949 | 566 | 271 | 7112 |
| 10-April | 9426 | 665 | 332 | 8429 |
| 11-April | 11177 | 781 | 406 | 9990 |
| 12-April | 13254 | 917 | 497 | 11840 |
| 13-April | 15716 | 1077 | 608 | 14031 |
| 14-April | 18636 | 1265 | 743 | 16627 |
| 15-April | 22098 | 1486 | 909 | 19702 |
| 16-April | 26203 | 1746 | 1113 | 23345 |
| 17-April | 31071 | 2050 | 1361 | 27659 |
| 18-April | 36843 | 2408 | 1666 | 32769 |
| 19-April | 43688 | 2829 | 2038 | 38821 |
| 20-April | 51804 | 3323 | 2493 | 45988 |
| 21-April | 61428 | 3903 | 3050 | 54475 |
| 22-April | 72840 | 4584 | 3732 | 64524 |
| 23-April | 86373 | 5384 | 4566 | 76422 |
| 24-April | 102419 | 6324 | 5587 | 90508 |
| 25-April | 121446 | 7428 | 6835 | 107183 |
| 26-April | 144008 | 8724 | 8363 | 126921 |
| 27-April | 170762 | 10247 | 10232 | 150283 |
| 28-April | 202486 | 12036 | 12518 | 177931 |
| 29-April | 240103 | 14137 | 15316 | 210650 |
| **30-April** | **284709** | **16605** | **18739** | **249365** |

**Table 2.** Predictions based on the mathematical model for reproduction number until April 30,2020

|  |  |  |  |
| --- | --- | --- | --- |
| 04-Apr | 459 | 17-Apr | 7456 |
| 05-Apr | 569 | 18-Apr | 9235 |
| 06-Apr | 706 | 19-Apr | 11437 |
| 07-Apr | 875 | 20-Apr | 14165 |
| 08-Apr | 1085 | 21-Apr | 17543 |
| 09-Apr | 1344 | 22-Apr | 21726 |
| 10-Apr | 1666 | 23-Apr | 26906 |
| 11-Apr | 2064 | 24-Apr | 33321 |
| 12-Apr | 2557 | 25-Apr | 41266 |
| 13-Apr | 3168 | 26-Apr | 51105 |
| 14-Apr | 3924 | 27-Apr | 63289 |
| 15-Apr | 4860 | 28-Apr | 78377 |
| 16-Apr | 6020 | 29-Apr | 97063 |
|  |  | 30-Apr | 120203 |

**Table 3.** Availability of Public health facilities in India

| Table 3. Availability of Public health facilities in India |  |
| --- | --- |
| Total number of govt. Hospitals | 23582 |
| Total No. of beds available | 710761 |
| Population coverage by one hospital | 56522 |
| Population coverage by one bed in a hospital | 1875 |
| No. of COVID-19 cases per Hospital (projected by exponential growth model) | 11 |
| No. of COVID-19 cases per Hospital ( projected by mathematical model on Ro) | 5 |

**Figure 2.**
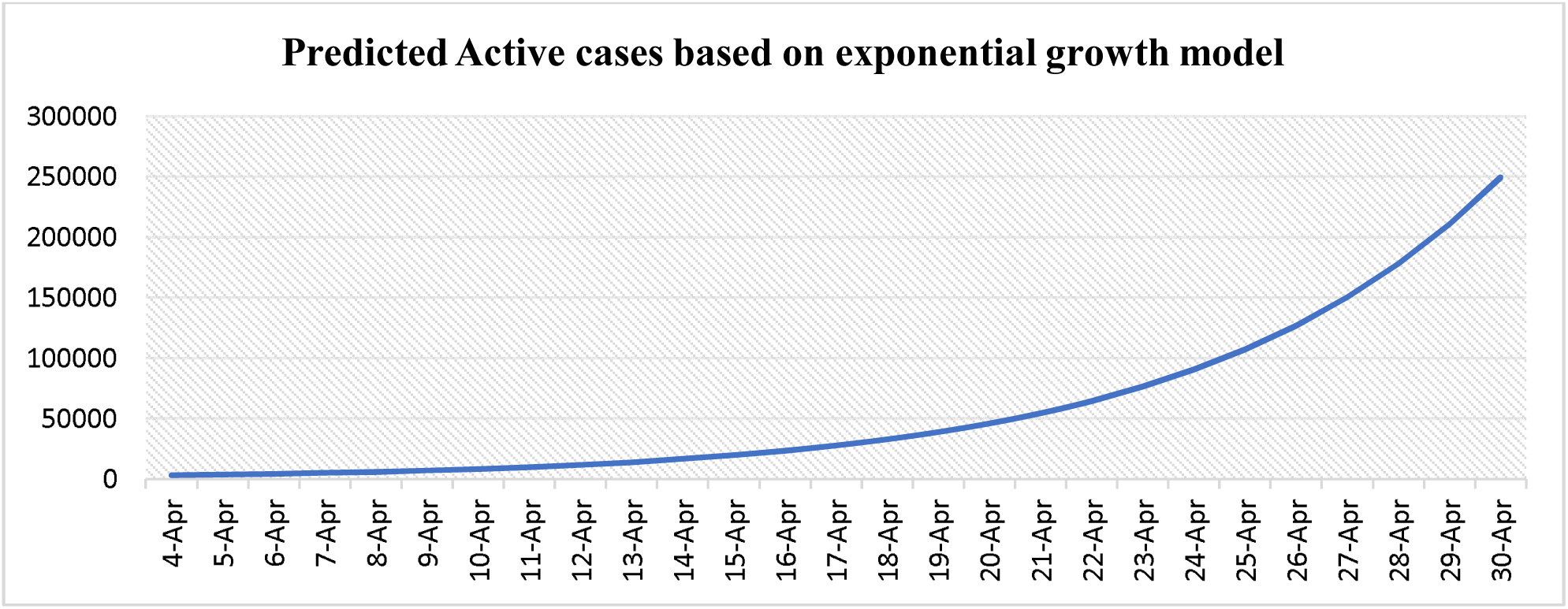
Predicted active cases based on exponential growth model for COVID-19 in India from the 4th of April till the 30th of April

The reproduction number for India comes out to be 2.56 based on the reported cases in 21 days which indicates one infected individual has infected averagely 2.56 individuals in this period of 21 days. The reproduction number may decrease in the coming time for India due to the preventive measures taken by government. The Herd Immunity for COVID-19 in India is estimated as 61% indicating that if atleast 61% of susceptible population has immunization to COVID-19, it can result in the elimination of infection from the population. The number of cumulative cases if predicted by the mathematical model for reproduction number comes out to be 1,20,203 until 30^th^ of April.

Total number of government hospitals functioning in India is 23,582 and total number of beds available in those hospitals is 7,10,761. If we consider the total population of India, the burden on public health facilities is quite high. One government hospital covers an average total population of 56,522 and there is one government hospital bed for a population of 1875. There will be around 11 COVID-19 cases in one government hospital if total number of cases are projected by exponential growth model and around 5 COVID-19 cases if cases are projected by mathematical model by 30^th^ of April. This results indicate that the public hospitals should be prepared in terms of isolated wards, beds and ICU’s.

## Discussion

Though this growth model provides the crude predictions for COVID-19 in India, but still provides an alarming situation for India in terms of public health preparedness. The active cases will require hospitalizations and ICUs in severe cases. If the exponential growth in the number of confirmed cases continues to grow in upcoming days, this may result in a public health disaster that is being already seen in several countries affected by COVID-19. Therefore, it may not be unrealistic to assume that what has happened in those countries can also happen in India. The density of the health workforce of India is among the lowest. The density of Physicians (7.8 per 10000 population) and nurses (21.1 per 10000 population) is low as compared to the world’s average.^20^ As compared to developed countries’ nurse-to-physician ratio of 3:1, India’s ratio is only around 0.6:1. This is a severe issue as the majority of them are concentrated in urban areas catering to only 20% of India’s population.^21^ depicting this public health capacity of India, it’s a matter of grave concern to address the need for hospitalization and ICUs in public health facilities in the case of the sudden outbreak of COVID-19 in India.

In the case of India, there is a serious concern about the number of tests done to detect the COVID-19. In that case, the number of confirmed cases can be seen as just the tip of an iceberg. As per the reports from the Indian Council of Medical Research (ICMR) total of 69,245 samples have been tested till the 3^rd^ of April., which is very less as compared to the tests done in other countries. As the number of Operational Government laboratories has been increased to diagnose the COVID-19 in India, it is expected that the number of testings will increase in the upcoming days, which will ultimately result in an increase in the number of confirmed cases.

This prediction has critical importance as it will not only help the government to have an estimate of the number of cases in the near future but also help them to plan the required strategy to accomplish the requirement in the given time to avoid any unfavorable condition. Although the Indian government has taken many preventive measures such as complete lockdown for three weeks, national and international travel restrictions, compulsory quartine, etc. to suppress the spread of disease, yet there may be sudden outbreaks within the communities which will end up in piling up the burden that is already there.

## Data Availability

Data has been collected from the data-sharing portal www.covid19india.org for India.

